# Investigating lesion location in relation to cerebellar mutism following pediatric tumor resection

**DOI:** 10.1101/2023.01.12.23284375

**Authors:** Jax Skye, Joel Bruss, Sebastian Toescu, Kristian Aquilina, Gino Bardi Lola, Aaron D. Boes

## Abstract

**Background and Objectives:** Approximately 25% of pediatric patients who undergo cerebellar tumor resection develop cerebellar mutism syndrome (CMS). Our group recently showed that damage to the cerebellar outflow pathway is associated with increased risk of CMS. Here, we tested whether these findings replicate in an independent cohort.

**Methods:** We evaluated the relationship between lesion location and the development of CMS in an observational study of 56 pediatric patients who underwent cerebellar tumor resection. We hypothesized that individuals that developed CMS after surgery (CMS+), relative to those that did not (CMS-) would have lesions that preferentially intersected with: 1) the cerebellar outflow pathway, and 2) a previously generated ‘lesion-symptom map’ of CMS. Analyses were conducted in accordance with pre-registered hypotheses and analytic methods (https://osf.io/r8yjv/).

**Results:** We found supporting evidence for both hypotheses. Compared with CMS-patients, CMS+ patients (n=10) had lesions with greater overlap with the cerebellar outflow pathway (Cohen’s d=.73, p=.05), and the CMS lesion-symptom map (Cohen’s d=1.1, p=.004).

**Discussion:** These results strengthen the association of lesion location with risk of developing CMS and demonstrate generalizability across cohorts. These findings may help to inform the optimal surgical approach to pediatric cerebellar tumors.

## Introduction

The odds of survival from pediatric brain tumors have steadily improved in recent decades,^1^ escalating the importance of understanding and preventing long-term treatment-related adverse effects. Approximately 25% of children undergoing cerebellar tumor resection will experience a postoperative syndrome characterized by emotional lability, executive dysfunction, and language deficits.^2-8^ This constellation of symptoms is commonly referred to as cerebellar mutism syndrome (CMS),^9^ posterior fossa syndrome, or cerebellar cognitive affective syndrome +/- cerebellar mutism; we will use the term CMS for this paper. The duration and severity of this syndrome is variable, but importantly, patients who develop CMS typically have worse long-term cognitive outcomes,^5, 10-13^ highlighting the importance of this syndrome as a potential harbinger of long-term impairment.

The pathophysiology of CMS is not fully understood. Recent work by our group has demonstrated the importance of lesion location. We found support for the hypothesis that damage to the cerebellar outflow pathway, an efferent pathway passing from the deep cerebellar nuclei through the superior cerebellar peduncles to the thalamus, is associated with the development of CMS in pediatric patients.^7^ We proposed this is likely due to an anatomical bottleneck whereby lesions to the cerebellar outflow pathway can disrupt communication of the cerebellum with a wide array of forebrain regions. This observation is consistent with findings from prior studies that have studied the role of lesion location as a risk factor for CMS.^14-25^

Identifying lesion sites associated with a higher risk of CMS has the potential to inform clinical practice. It is possible that more personalized prognostic information could be provided to patients about their risk for CMS based on the tumor location relative to the cerebellar outflow pathway. It is also possible that once critical anatomical regions associated with CMS are well established they could be identified in advance of the tumor resection and used to inform imaging-guided surgery to avoid those regions when possible. However, an important prerequisite to the development of any clinical application is demonstrating that lesion location is a reliable marker of risk for CMS that generalizes to other independent cohorts. This was the primary objective of the current study. Specifically, we aimed to evaluate whether the same anatomical regions associated with higher risk of CMS in our previous study would replicate in an independent cohort. Our anatomical hypotheses and the analytic strategy were pre-registered with the Open Science Framework (https://osf.io/r8yjv/).

In a cohort of 56 patients who underwent cerebellar tumor resection, we hypothesized that patients who developed CMS (CMS+) would have damage to the cerebellar outflow pathway to a greater extent than individuals who did not develop CMS (CMS-). Similarly, we hypothesized that CMS+ individuals would have lesions that overlapped with a lesion-symptom ‘map’ of CMS previously derived from a sample of 195 patients.^7^

## Methods

We analyzed clinical outcome and imaging data in patients with cerebellar tumor resection surgery (n=56) from two sites: University of Iowa (n=9) and Great Ormond Street Children’s Hospital in London (n=47).^7^ We included patients under the age of 21 with a diagnosis of a cerebellar tumor that had a surgical resection and follow-up imaging to show the tumor resection cavity. Each patient also had clinical assessments by their treatment teams to determine whether they met criteria for CMS. CMS was defined by criteria outlined previously: post-surgical onset of reduced speech\mutism and emotional lability. Additional common features included motor dysfunction or hypotonia.^9^ Post-surgical structural neuroimaging was conducted at least one month from the date of tumor resection. All scans were reviewed in advance of the analysis and only included if they were of sufficient quality to clearly observe the borders of the post-surgical resection cavity. Scans were performed for clinical indications, so scanners and MRI sequence varied between patients. The boundaries of each lesion were manually traced on the patient’s T1 native scan before being transformed to MNI152 space using ANTs.^26^ The anatomical accuracy of the lesion tracing and the transformation to MNI space was confirmed by a neurologist (A.D.B.) blinded to CMS status.

Our first hypothesis was that CMS+ patients would have greater disruption to the cerebellar outflow pathway. The region of interest that defines the cerebellar outflow pathway was the same as previously described.^7^ Briefly, it was produced by combining the cerebellar deep nuclei atlas (Spatially Unbiased Infra-Tentorial Template, diedrichsenlab.org/imaging/propatlas.htm) with a mask of the superior cerebellar peduncles defined from a probabilistic atlas of these tracts.^27^ The extent to which each individual patient’s lesion location, or lesion ‘mask’, intersected with the binary cerebellar outflow pathway ROI was quantified and referred to as cerebellar outflow pathway ‘lesion load’. The percentage of voxels in each slice of the cerebellar outflow pathway that intersected with the lesion mask was calculated. This required first separating the cerebellar outflow pathway ROI by 1mm oblique (17°) coronal slices perpendicular to the superior cerebellar peduncles. The slice of the cerebellar outflow pathway with the highest percentage of overlap between the lesion mask and ROI was used as the value of cerebellar outflow pathway ‘lesion load’ for that patient. A lesion intersecting all cerebellar outflow voxels would have a value of 100% and a lesion that entirely spares this outflow pathway would have a value of 0%. The lesion load values of the CMS+ and CMS-groups were compared using an independent samples *t*-test to evaluate the one-tailed hypothesis that higher cerebellar outflow pathway lesion load will be seen in the CMS+ group compared with CMS-group.

This study’s second main hypothesis was similar in design to the first but used a data-driven *a priori* ROI in place of the cerebellar outflow pathway ROI. We used the lesion-symptom map that was generated by Albazron and colleagues^7^ using lesion and outcome data from 195 pediatric patients. The multivariate lesion-symptom mapping was performed using the R package LESYMAP.^28^ LESYMAP uses sparse canonical correlation analysis for neuroimaging (SCCAN) to associate loci of brain damage with CMS status as a binary outcome. A within-sample cross validation is performed with mapping in 75% of the patients to predict the CMS status of the remaining 25%. This determines the optimal sparseness value with the highest cross-validation correlation between the measured and predicted score.^28^ The resulting lesion-symptom map identified cerebellar regions statistically associated with severe post-operative cognitive and affective symptoms, which were referred to as cerebellar cognitive affective syndrome in that study, but also met the criteria for CMS as used in the current analysis. This statistical map showed localization to the cerebellar outflow pathway, specifically the fastigial nuclei, interposed and medial dentate nuclei, superior cerebellar peduncles and also regions outside of the cerebellar outflow pathway, including lobules IX and X of the vermis.^7^ Using this lesion-symptom map as an *a priori* ROI we tested our second hypothesis that CMS+ patients will have a higher lesion-symptom map lesion load than patients without CMS. We created lesion load values for each patient by summating the voxels of the lesion-symptom map intersecting with each individual’s lesion mask. We compared the lesion-symptom map lesion load between the CMS+ and CMS- groups, again using a one-tailed independent samples *t*-test.

### Data Availability and Access

The principal author takes full responsibility for the data, analyses, interpretation and conduct of research; has full access to all the data; and has the right to publish any and all data separate and apart from the guidance of any sponsor.

### Standard Protocol Approvals, Registrations, and Patient Consents

This study was approved by the Institutional Review Board of each institution. Approval from an ethical standards committee to conduct this retrospective study was received.

## Results

A total of 56 pediatric patients who underwent posterior fossa tumor resections met inclusion criteria for the study. 10 patients had CMS (18%). The average age of the sample 6.2 ± 3.7 years, median 6.0 years, range 5 months – 14 years with various types of tumors (24 pilocytic astrocytomas, 24 medulloblastomas, 3 atypical teratoid rhabdoid tumors, 2 gangliomas, 1 ependymoma, 1 hemangioblastoma, and 1 high-grade glioma). Children with CMS were younger (x□_CMS+_=4.4 years old ± 2.4 years, x□_CMS-_=6.9 years old ± 4.2 years; t(21)=-2.59, p=.016) and had similar lesion volume (x□_CMS+_=6076 mm^3^ ± 4891 mm^3^, x□_CMS-_=9831 mm^3^ ± 10742 mm^3^; t(30)=-1.7, p=.10). The lesions of all CMS+ patients crossed the midline and 65% of CMS-lesions crossed the midline. The lesion masks from each patient were overlapped to show the spatial distribution of the lesions in the entire sample (Figure 1A) and split by CMS status. CMS+ individuals (n=10) are shown in Figure 1B and CMS-individuals (n=46) are shown in Figure 1C. A proportional subtraction map of CMS+ minus CMS-lesion masks is displayed in Figure 1D.^29^

**Figure 1.**
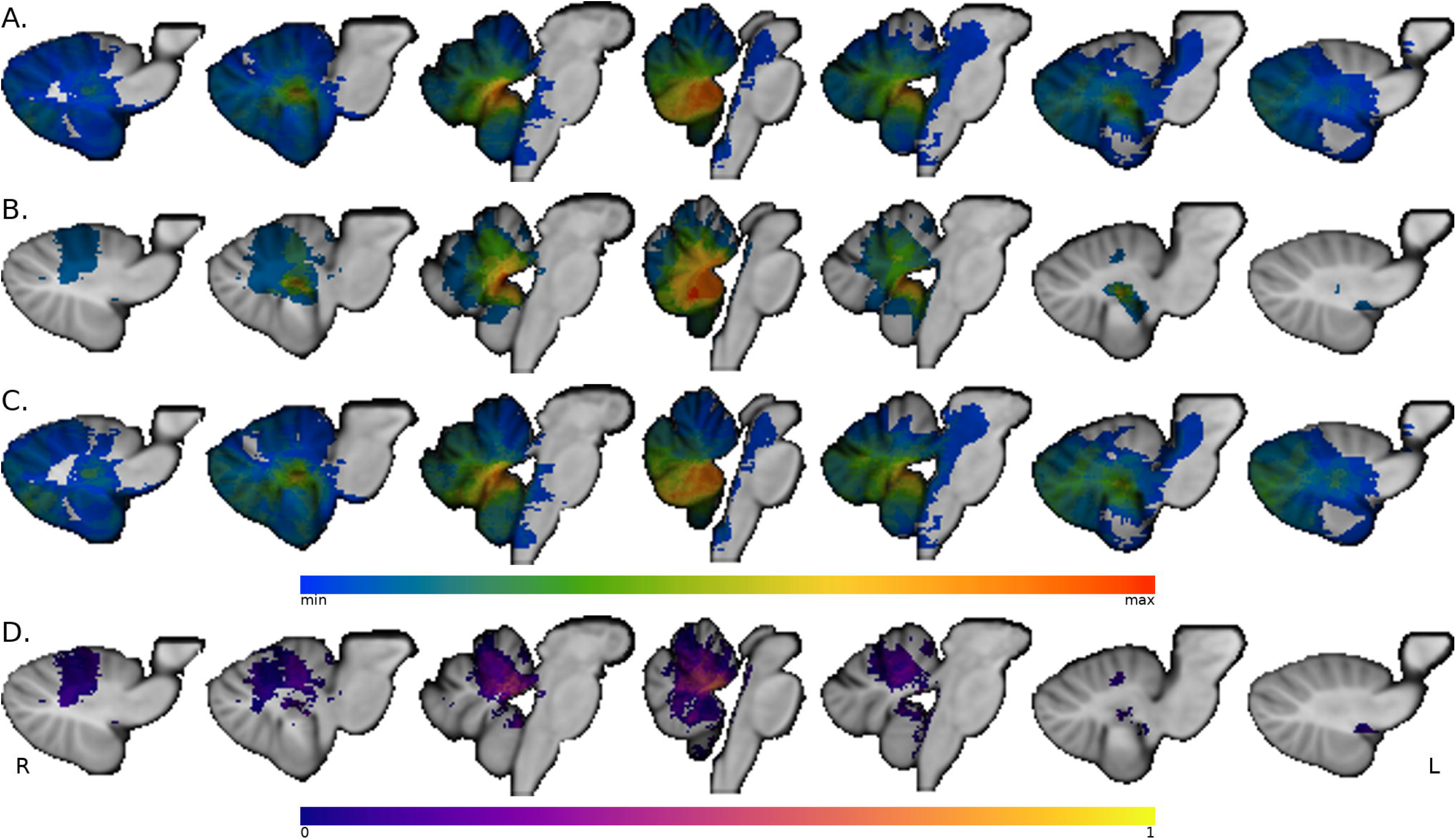
A – D. 1A shows the lesion overlap of all patients included in this study (n=56). The region of maximum lesion overlap (n=29 of 56) was the right vermian lobule IX (MNI coordinates 4 -53 -34). 1B shows the CMS+ lesion overlap (n=10) with the maximum overlap at the right vermian lobule IX (9 of 10 lesions, MNI 3 -54 -34). 1C displays the CMS-lesion overlap with peak overlap of 21 of 46 lesions in the right vermian lobule VIII at MNI coordinate 2 -61 -36. 1D shows the proportional subtraction map with a regional peak in the anterior vermis at MNI coordinate 3 -47 -26.

To test our *a priori* hypothesis that damage to the cerebellar outflow pathway (Figure 2A) increases the risk of developing CMS, cerebellar outflow pathway lesion load values were calculated for patients with and without CMS. The cerebellar outflow pathway lesion load was higher in the CMS+ group relative to the CMS- group (x□_CMS+_=37%, *σ*_CMS+_=30% and x□_CMS-_ =19%, *σ*_CMS-_=24%, respectively; t(11)=1.8, p=.050; Cohen’s d=0.73). Notably, the CMS rate was observed to increase in accordance with greater lesion involvement of the cerebellar outflow pathway (Figure 3). We evaluated our second hypothesis that CMS+ patients would have a greater lesion-symptom map lesion load when compared to CMS-patients (lesion-symptom map shown in Figure 2B). As hypothesized, CMS+ patients had a greater lesion-symptom map lesion load than CMS- patients (x□_CMS+_=.018 ± .010 and x□_CMS-_=.0090 ± .011, respectively; t(14)=2.4, p=.014; Cohen’s d=0.79).

**Figure 2.**
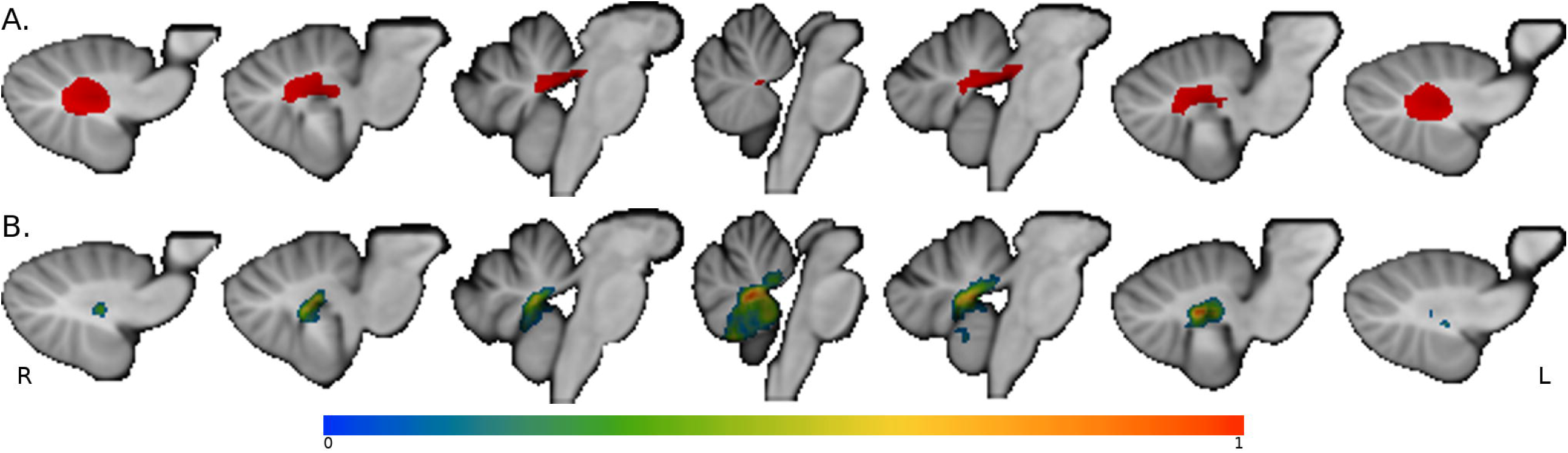
A&B. 2A shows the cerebellar outflow pathway in red. 2B shows the lesion-symptom map from Albazron and colleague’s 2019 study used to generate lesion-symptom map lesion load.

**Figure 3.**
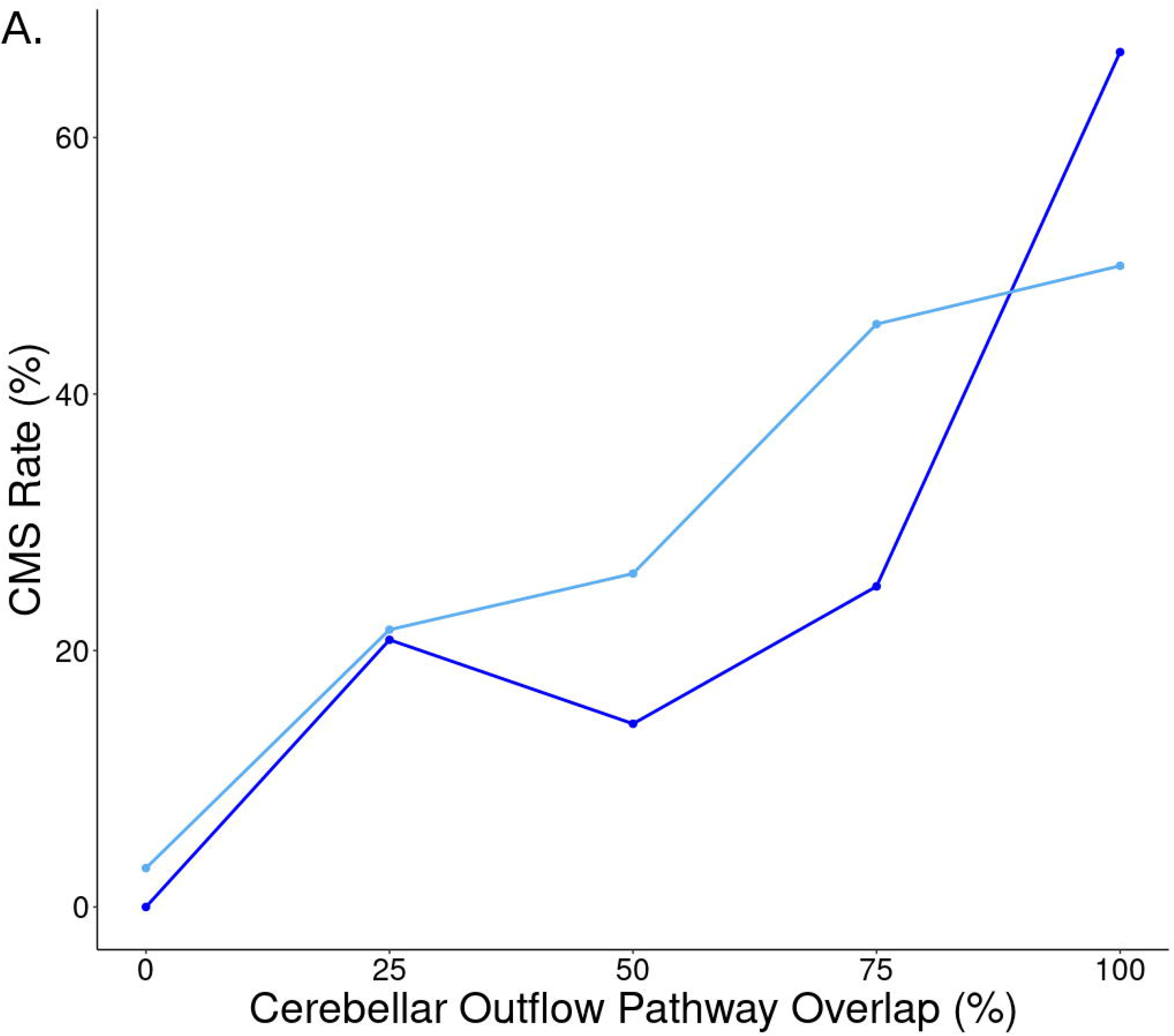
The degree of overlap between a patient’s lesion and the cerebellar outflow pathway is systematically associated with their likelihood of developing CMS. The rate of CMS development as a function of cerebellar outflow pathway overlap in the current study (N=56) is shown in dark blue; the lesion of every CMS+ patient intersects with the cerebellar outflow pathway. The same relationship for patients in the Albazron study (N=195) is shown in light blue.

## Discussion

This study evaluates lesion location in relation to developing cerebellar mutism syndrome after cerebellar tumor resection in pediatric patients.^5-7^ Our results support prior work in showing that lesion location is associated with risk of CMS. We tested two pre-registered hypotheses and found supporting evidence for both. Individuals that developed CMS had lesions with greater overlap with the cerebellar outflow pathway than those that did not develop CMS. In addition, patients with CMS had lesions that overlapped to a greater extent with a lesion-symptom map of CMS derived from an independent sample.^7^ Taken together, these findings emphasize that lesion location is useful in predicting the development of CMS in a way that generalizes across cohorts.

There are limitations of this study. First, CMS was diagnosed by the treating clinicians without any formal assessments of behavioral deficits. It is possible that symptom-specific quantitative assessments may further clarify unique anatomical associations with specific symptoms. Other factors that likely influence the development of CMS, like post-surgical treatment plan, surgical approach, edema, hydrocephalus, and premorbid cognitive abilities were not evaluated here. The observation that lesion location significantly relates to CMS outcome without accounting for these other variables supports the robust effect of lesion location, but more sophisticated models that take these additional factors into account are likely to be most useful in assessing CMS risk.

In closing, this study provides additional evidence that damage to critical regions of the cerebellum and its outflow pathway is associated with increased risk of a child developing CMS after cerebellar tumor resection. Further work in this line of research could be used to inform the surgical approach to pediatric cerebellar tumor resection. For instance, critical anatomical regions that, when resected, are associated with increased CMS risk could be displayed as an overlay onto a patient’s pre-operative MRI scan so that surgeons could design a minimally invasive MRI-guided approach that minimizes damage to these regions.

## Data Availability

All data produced in the present study are available upon reasonable request to the authors.

## Glossary

CMS: cerebellar mutism syndrome
CMS+: individuals with cerebellar mutism syndrome
CMS: individuals without cerebellar mutism syndrome

